# Molecular phenotypes associated with antipsychotic drugs in the human caudate nucleus

**DOI:** 10.1101/2021.10.11.21264848

**Authors:** Kira A. Perzel Mandell, Nicholas J. Eagles, Amy Deep-Soboslay, Ran Tao, Shizhong Han, Richard Wilton, Alexander S. Szalay, Thomas M. Hyde, Joel E. Kleinman, Andrew E. Jaffe, Daniel R. Weinberger

## Abstract

Antipsychotic drugs are the current first-line of treatment for schizophrenia and other psychotic conditions. However, their molecular effects on the human brain are poorly studied, due to difficulty of tissue access and confounders associated with disease status. Here we examine differences in gene expression and DNA methylation associated with positive antipsychotic drug toxicology status in the human caudate nucleus. We find no genome-wide significant differences in DNA methylation, but abundant differences in gene expression. These gene expression differences are overall quite similar to gene expression differences between schizophrenia cases and controls. Interestingly, gene expression differences based on antipsychotic toxicology are different between brain regions, potentially due to affected cell type differences. We finally assess similarities with effects in a mouse model, which finds some overlapping effects but many differences as well. As a first look at the molecular effects of antipsychotics in the human brain, the lack of epigenetic effects is unexpected, possibly because long term treatment effects may be relatively stable for extended periods.

## Introduction

Schizophrenia is a serious mental illness which is characterized by psychosis as well as other symptoms that disrupt cognitive and social functioning. Antipsychotic drugs are a common first line treatment for schizophrenia and many other psychotic conditions. Their mechanism of action has been linked primarily with antagonism of dopamine type II receptors ^1^ but other neurotransmitter receptors are involved in the actions of several of these agents. It is noteworthy, however, that these drugs are imperfect and also known to cause a wide array of neurologic and metabolic side effects, which have driven a mission to create more effective drugs with fewer off-target side effects ^2^. A challenge in the pursuit of better antipsychotic treatment has been an overall poor understanding of the molecular underpinnings of the disease. While examining the effects of antipsychotics may not elucidate some of the causative mechanisms of the illness, it might identify mechanisms that are critical for effective treatment, and can further help partition and interpret case-control associations in the context of potentially causal versus consequential effects.

Two important molecular substrates for potentially capturing cellular effects of various pharmacological interventions are gene expression and DNA methylation (DNAm). DNAm is an epigenetic regulator of gene expression. It occurs most commonly at CpG dinucleotides, but in neurons also uniquely occurs at CpH sites (H = A, T, or C). It is thought to be a reflection of the interaction between genes and environment, as various environmental factors including diet ^3^ and cigarette smoking ^4^ have been associated with altered methylation patterns at specific sites in the genome. Thus, DNA methylation analysis has the potential to impart the effects of drugs such as antipsychotics on the epigenome.

Previous studies have aimed to uncover the molecular footprint of antipsychotic effects in a variety of ways. In humans, most studies examining molecular effects of antipsychotics have been performed in peripheral tissues such as blood ^5–7^ and non-CNS tissues like adipose ^8^. These studies have provided some insight into how antipsychotics may alter DNAm and gene expression levels, but it is unclear to what extent these effects are present in brain. Additionally, these DNAm studies used microarray technology, which only captures a small fraction of CpG dinucleotides in the genome and does not target CpH stes. Studies that have been performed in human postmortem brain tissue of individuals with schizophrenia, many of whom had received antipsychotics, have identified many molecular associations with genetic risk ^9 10^, but have identified relatively few direct case-control differences in gene expression. Minimal differences in DNAm levels have been reported between cases and controls ^11^. Further, none of the prior studies of gene expression or DNAm in postmortem human brain tissue from donors with schizophrenia have investigated specifically the effects of antipsychotic use. The previous literature contains many studies that examined antipsychotic-induced differences in brains of model organisms such as mice ^12^, rats ^13^ and rhesus monkey ^14^, but how well these findings translate to the human brain and clinical treatment is unclear.

The caudate nucleus is especially highly involved in dopaminergic signaling, with involvement in motor, learning, and reward processes. PET studies of dopamine (DA) activity and DRD2 availability in patients with schizophrenia have highlighted the caudate nucleus as the site of primary DA relevance to illness and treatment ^15^. While many postmortem human brain studies of gene expression in schizophrenia have examined the prefrontal cortex, a region prominently linked with the so-called negative and cognitive features of schizophrenia, its role in the treatment effects of DRD2 antagonists is uncertain and DA receptors in prefrontal cortex are at least one order of magnitude less abundant than in caudate ^16^. A recent study of gene expression and schizophrenia genetic risk in caudate has identified far more differential expression by disease status than other brain regions (like hippocampus and frontal cortex), highlighting the need for further investigation of this region ^17^. Here we examine associations of gene expression and DNA methylation levels in the human postmortem caudate nucleus to antipsychotic treatment to better characterize their molecular landscapes.

## Results

### Gene expression associations to antipsychotics in the human caudate nucleus

In order to assess the molecular consequences of antipsychotic use in the human brain, we first analyzed RNA-seq data from the caudate nucleus of 380 postmortem brains, including samples from 147 patients diagnosed with schizophrenia and 233 adult neurotypical controls; (see Methods) ^17^. We used postmortem toxicology assays measured at time of death to classify the 147 patients into 100 who were antipsychotic positive at time of death (noted as SCZDAP), and 47 who were antipsychotic negative by toxicology (noted as SCZD). Antipsychotic toxicology testing was chosen for analysis (as opposed to reported use via next-of-kin or medical records) to ensure that donors were indeed compliant with their antipsychotic prescriptions at time of death. Almost all schizophrenia patients used antipsychotics at some point in life. Demographics were largely consistent among these three groups (Table 1).

**Table 1:** Demographics of sample groups. For sex, M denotes males and the remaining proportions of samples are female. For race, AA denotes African American ancestry, and the remaining proportions of samples are of European ancestry. For instances where patient group demographics are significantly different from control demographics, significance is noted as: p < 0.001 ***, p < 0.01 **, p < 0.05 *

|  | Control | SCZD | SCZDAP |
| --- | --- | --- | --- |
| n | 233 | 47 | 100 |
| Lifetime Antipsychotic | 0% | 96% *** | 100% *** |
| Sex | 72% M | 70% M | 68% M |
| Age range (median) | 17-89 (49) | 17-96 (56) * | 18-81 (51) |
| Race | 50% AA | 55% AA | 54% AA |

We performed a series of regression analyses to refine the relationship between antipsychotic status, schizophrenia diagnosis and gene expression, including defining (1) schizophrenia effects ignoring APs (147 schizophrenia cases versus 233 controls), (2) AP effects contrasting patient groups (100 SCZDAP versus 47 SCZD) and (3) AP effects independent of diagnosis (100 SCZDAP versus 280 SCZD + Control). All three analyses were performed on the same set of 380 samples.

For a frame of reference, we first performed linear modeling to identify genes which were differentially expressed based on schizophrenia diagnosis. We identified 3131 significantly differential genes at FDR < 0.05 (Table S1). Our findings are in line with previous similar analysis by Benjamin et al. ^17^

Next, we performed linear modeling to determine which genes were differentially expressed based on antipsychotic status while adjusting for clinical and technical confounders (see Methods). When assessing the differences between patient groups - SCZDAP and SCZD, we identified 70 genes that were significantly differentially expressed at FDR < 0.05 (Table S2, Figure 1A). Effect sizes were generally small, with a mean gene expression difference of 1.25% (log2 fold change = 0.32).

**Figure 1:**
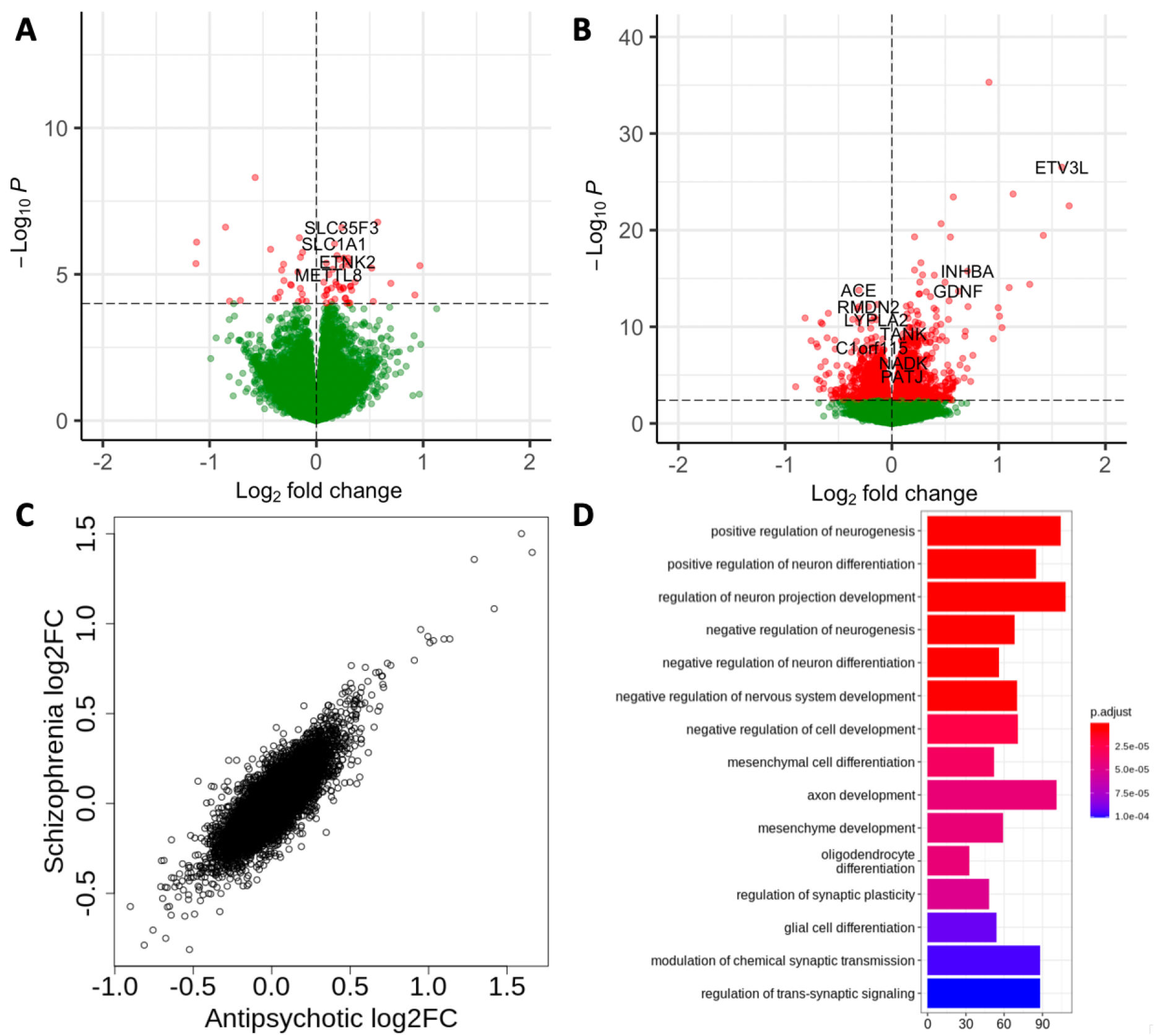
Gene expression differences by antipsychotic toxicology data. (A) Volcano plot of differentially expressed genes by antipsychotic toxicology when examining the contrasts between patients (SCZD vs SCZDAP). Points in red are genes that surpass FDR < 0.05 significance. (B) Volcano plot of differentially expressed genes by antipsychotic toxicology when examining all samples (SCZDAP vs SCZD + Control). Points in red are genes that surpass FDR < 0.05 significance. (C) Comparison of effect sizes on gene expression when modelling antipsychotic toxicology vs modelling diagnosis. Diagnosis and antipsychotic effects are overall highly similar. (D) Gene ontology (GO) terms for gene sets which are enriched among genes which are significantly differentially expressed by antipsychotic toxicology when examining all samples (SCZDAP vs SCZD + Control).

Because this sample size is relatively small and most SCZ patients are on antipsychotics, we expanded our analysis to include neurotypical controls. In this analysis, we compared SCZDAP patients to all samples who were not on antipsychotics - SCZD and controls. We identified 2347 genes differentially expressed between these groups (Table S3, Figure 1B). For the remainder of the results, this is the gene set that will be examined for antipsychotic differential expression. *DRD2*, a gene which has been shown to have increased expression in response to antipsychotics in animal studies, reaches near significance (P = 0.004, FDR = 0.0509) for increased expression with antipsychotics. This gene set was enriched for many gene ontology (GO) terms, with the enrichment for terms related to synaptic signaling and neurogenesis, processes which have been frequently implicated in the pathology of schizophrenia (Figure 1D, Table S4). Generally, the effects on differential expression by antipsychotics and disease status are very similar and highly correlated (Figure 1C), indicating that antipsychotic use may be driving broader differences that have been identified in prior studies between cases and controls. However, 26% of these genes were not differential between cases and controls, providing evidence that some of these differences may be specifically driven by antipsychotic use at least around the time of death. As with previous findings in case-control differences, effect sizes were subtle, with a mean gene expression difference of 1.12% (log2 fold change = 0.16). 57 genes had a greater difference (log2FC > 0.5, % change = 1.41%).

Another factor in assessing effects of antipsychotics is consideration of the differences between generations of the drugs - older “typical” antipsychotics (such as haloperidol and chlorpromazine) and newer “atypical” antipsychotics (including clozapine, olanzapine, and risperidone). Typicals are generally more selective as specific DRD2 antagonists, while atypicals target other receptor systems, particularly 5HT2. We performed several analyses to tease apart the effect differences of typicals and atypicals, but generally we found little to no significant difference between the groups. These analyses were complicated by the fact that some patients were on both generations of antipsychotics at once and that the subgroupings became increasingly underpowered.

### Lack of DNAm associations to antipsychotic use

DNA methylation (DNAm) is thought to be a molecular representation of environmental exposures, as well as an effector of gene expression, so we investigated antipsychotic effects on DNAm by examining whole genome bisulfite sequencing (WGBS) data from 296 postmortem caudate nuclei (121 cases, 175 controls). In this sample, 86 schizophenia patients were antipsychotic positive at time of death as determined by toxicology assays, leaving 35 patients who were not. The vast majority of these samples (291, 98.3%) of these samples were also included in the previous RNA-seq analyses, and both DNA and RNA were concurrently extracted from the same tissue aliquot. With WGBS, we modeled differences across 27,812,354 CpG sites and 50,336,332 CpH sites (see Methods). In these analyses, comparing differences within schizophrenia patients as well as including controls, we found no significant differences after adjusting for multiple testing, and overall, p-values were depleted for low values. Further, there was no significant difference in DNAm between the different generations of antipsychotics, and there was no significant difference in DNAm by diagnosis. This indicates that the observed differences in gene expression were not associated with differential DNAm, and challenges the idea that DNAm can represent acute differences in environmental exposures.

### Antipsychotic related changes are not consistent between brain regions

To understand how applicable our findings were to other regions of the brain, we performed the same analyses in the dorsolateral prefrontal cortex (DLPFC). Like the caudate, there were no significant differences in DNAm between those on antipsychotics at time of death and those who were not (N = 165).

When examining gene expression via RNA-seq, we found that overall there were fewer differences in the DLPFC than in caudate. The DLPFC sample included 117 cases and 197 controls, of which 82 were antipsychotic positive at time of death and 35 were antipsychotic negative. 235 of these samples came from donors who were also included in the caudate analyses. There were no significant differences when examining only SCZ cases grouped by toxicology status. When expanding the analysis to include controls as previously described, we identified 622 differentially expressed (FDR < 0.05) genes - far fewer than the 2347 identified in the caudate (Table S5). The sample sizes between these two brain regions were quite similar, so it is unlikely that this discrepancy can be attributed to differences in power.

Further, the genes which are differential in each region do not strongly overlap between regions. Only 93 (15%) of the genes differentially expressed based on toxicology status in DLPFC were also differentially expressed in caudate (Figure 2A). When examining genes which were differentially expressed in caudate, the majority (79%) do not replicate in DLPFC at p < 0.05 (Figure 2B). One potential reason for these differences is difference in affected cell type. Genes differentially expressed by antipsychotic use in caudate are most enriched for specificity to D1 dopaminoceptive neurons (OR=3.50, P = 1.74e-82), D2 dopaminoceptive neurons (OR = 3.39, P=1.59e-77), and oligodendrocytes (OR=3.24,P = 7.84e-71). In DLPFC, they are most enriched for specificity to macrophages (OR= 3.69, P=8.70e-30), microglia (OR=7.22, P=6.99e-84), and T-cells (OR =2.43, P=2.94e-12), and depleted for specificity to oligodendrocytes (OR = 0.35, P = 2.60e-6) ^18^ (see Methods, Table S6, Table S7). Altogether, these results indicate that the caudate nucleus is strongly affected by antipsychotic drugs at a molecular level, and that these drugs have different effects in different areas of the brain.

**Figure 2:**
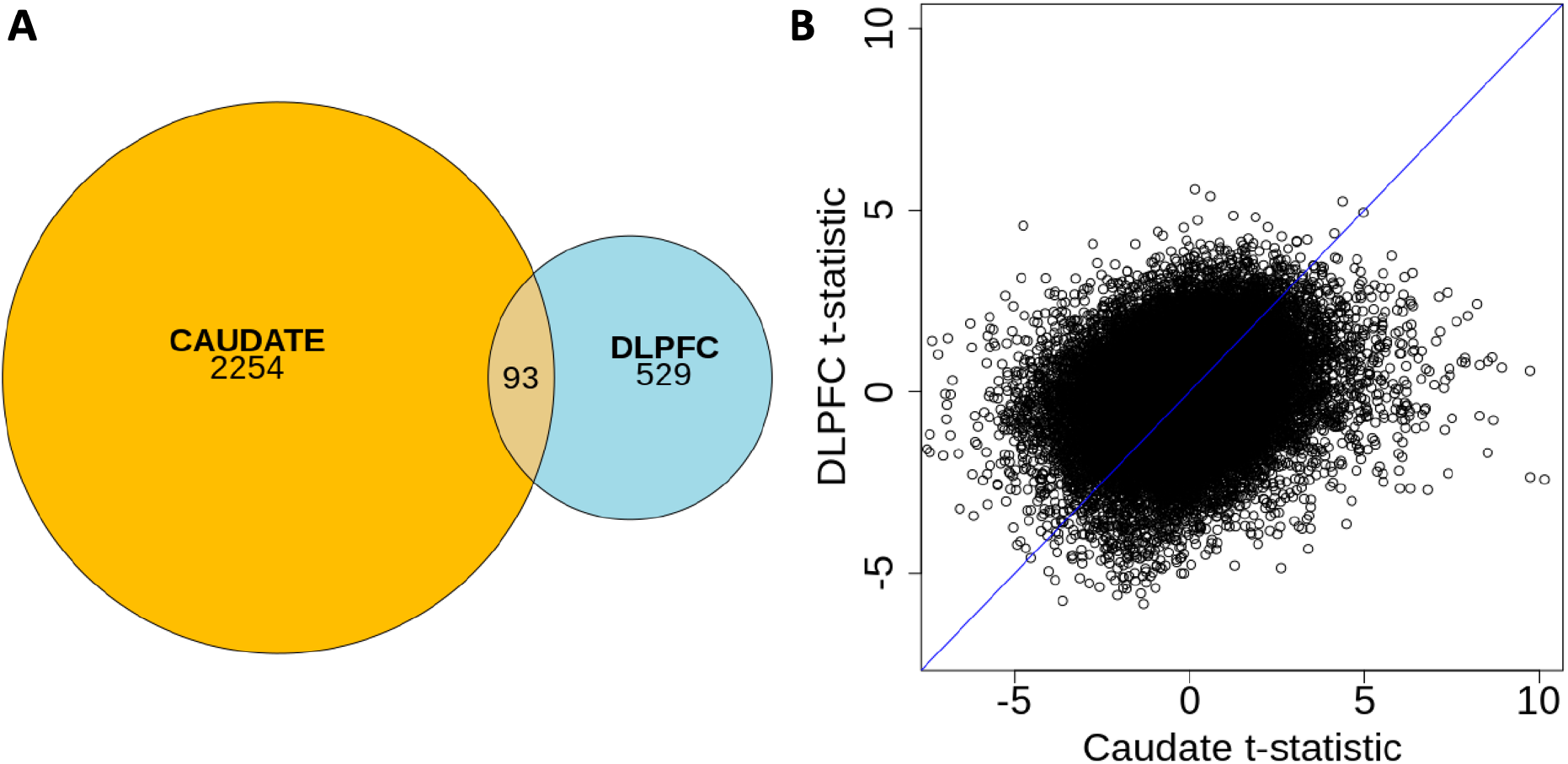
Comparison of antipsychotic-based differential gene expression in the caudate nucleus and the DLPFC. (A) A Venn diagram showing the relative proportions of differential gene expression in each region and highlighting the low overlap among genes which pass FDR < 0.05 in each region. (B) Comparison of antipsychotic association t-statistics for each gene in the caudate and in the DLPFC. We see that effects in each brain region are dissimilar. Pearson’s correlation of these values is r = 0.25.

### Translational considerations for mouse and human brain studies

A major difficulty in disambiguating antipsychotic and schizophrenia effects on the human brain is the fact that virtually all schizophrenia patients are treated with antipsychotics at some time in their history making molecular differences hard to tease apart. The toxicology screens are sensitive and accurate for documenting drug in brain at the time of death, but they do not provide a historical reference for prior treatment. For this reason, animal models are a potentially valuable tool in understanding the molecular effects of antipsychotics in the brain. We aimed to assess the translationability of such work by comparing our findings to results from Kim and colleagues ^12^, who investigated gene expression changes in mice treated with haloperidol - a typical antipsychotic. We found that genes that were significantly differentially expressed by antipsychotic use in human caudate were enriched for being differentially expressed in mouse (OR = 2.13, P = 0.01). The 17 genes that were significantly differentially expressed in both mouse and human were *LAMB3, EPHA4, ANXA3, FAT2, DOCK4, PENK, HECTD2, GDPD5, ANO2, HTR2A, REM2, HBA2, GAN, CHD3, CBLN4, CSTB*, and *PFKL*. Further, if we only looked at mouse hits from the striatum - a region that includes the caudate - we found slightly stronger enrichment (OR = 2.18, p = 0.01, see Methods).

We then assessed the replication of differentially expressed genes in mouse within our caudate dataset. We found that 34% of significant (q < 0.05) hits in mouse replicated in human caudate at p < 0.05. However, only 52% of the replicated genes are directionally consistent, and overall, effect sizes are very uncorrelated between mouse and human. These results indicate that there are some valuable similarities between antipsychotic effects in mice and human brains, but that there are differences which are important to understand as well.

## Discussion

Here we have examined the molecular effects of recent antipsychotic use in the human caudate nucleus. We found many changes of small effect in gene expression, which overlap highly with case-control differences, and a surprising lack of differences in DNAm levels. We also see that these effects are variable between brain regions and when contrasted with mice. It is interesting to note that genes differentially expressed in caudate based on antipsychotics presence at death highlighted synaptic signaling while in DLPFC it emphasized microglia and other immune cells. While these cell phenotypes in DLPFC have not been implicated in bioinformatic translation of GWAS risk genes, they have been found in differentially expressed gene sets comparing patients with schizophrenia to neurotypicals, suggesting that these prior case-control findings represent at least in large part antipsychotic treatment epiphenomena. Overall, our data provide an early view of antipsychotic effects in human brain, but the tip of the iceberg that we see is not likely to be the whole story.

There are many challenges to studying antipsychotic use in human brain. At a phenotypic level, we examined antipsychotic status at time of death, but nearly all schizophrenia patients have used antipsychotics at some point, and often for prolonged periods of time, throughout their life. Thus, we are only capturing evidence of acute antipsychotic treatment at a relatively restricted period of time. Longer term effects would have been masked here, and this may be the reason we see mostly subtle gene expression differences and virtually no DNA methylation differences. DNAm may more accurately represent long term, cumulative effects, rather than acute environmental changes. This may challenge notions about the plasticity of DNAm in response to acute exogenous factors.

Another difficulty in studying human postmortem brain in this particular study is that the samples used were bulk tissue - a mix of cell types. This could mask cell-type specific effects, and could also contribute to the difficulty of comparing results between different brain regions and organisms. The increasing presence of single-cell datasets will help elucidate whether antipsychotic-induced changes are cell-type specific, and whether these changes are consistent between different tissues and animals.

Overall, these findings provide a first look at the molecular genetic effects of antipsychotics in the human brain. It is clear that further investigation is warranted - especially to understand the translatability of tissue and animal studies.

## Methods

### Study samples

Details regarding postmortem human brain collection were described in a previous manuscript ^17^. Briefly, human brain tissue was obtained from two phases of collection. A large number of samples were obtained at the Clinical Brain Disorders Branch (CBDB) at National Institute of Mental Health (NIMH) from the Offices of the Chief Medical Examiner of Northern Virginia, Northern District and the District of Columbia Medical Examiners’ Office, according to NIH Institutional Review Board guidelines (Protocol #90-M-0142), with informed consent of legal next-of-kin. These samples were transferred to the Lieber Institute for Brain Development (LIBD) under a material transfer agreement with the NIMH. Additional samples were collected at the LIBD according to a protocol approved by the Institutional Review Board of the Maryland Department of Health (#12-24).

Retrospective clinical diagnostic reviews were conducted for every brain donor to include data from: autopsy reports, forensic investigations, neuropathological examinations, telephone screening, and psychiatric/substance abuse treatment record reviews and/or supplemental family informant interviews (whenever possible). All data was compiled and summarized in a detailed psychiatric narrative summary, and was reviewed independently by two board-certified psychiatrists in order to determine lifetime psychiatric diagnoses according to DSM-IV/V. For this study, 147 donors met criteria for schizophrenia, and 233 donors were free from all DSM-IV/V psychiatric/substance use disorder diagnoses. Non-psychiatric healthy controls were only included if free from all drugs of abuse by toxicology testing in blood, urine or brain at time of death.

Antipsychotic toxicology testing was initially performed at the medical examiner as part of routine toxicology testing during autopsy to determine cause of death. Additionally, supplemental toxicology was conducted for donors with schizophrenia, to screen for any prescribed medications being taken at time of death (at therapeutic levels) that medical records, medical examiner reports, or next-of-kin reported at time of death. Supplemental toxicology testing was completed at National Medical Services Laboratories in Horsham, PA (www.nmslabs.com), using postmortem blood or cerebellar tissue. Of the donors who were antipsychotic positive, 29% were on only typical antipsychotics, 56% were on only atypical antipsychotics, and 15% were on both types of antipsychotics at time of death.

The caudate nucleus was dissected from the slab containing the caudate and putamen at the level of the nucleus accumbens. The caudate was dissected from the dorsal third of the caudate nucleus, lateral to the lateral ventricle. DNA and RNA were concurrently extracted from ∼250mg of tissue using the QIAGEN AllPrep DNA/RNA Mini Kit.

### WGBS data generation

Extracted DNA was subjected to QC via Bioanalyzer and WGBS library construction using the Swift Accel-NGS Methyl-Seq DNA Library Kit (https://swiftbiosci.com/accel-ngs-methyl-seq-dna-library-kit/), with 0.1% Lambda spike-in (to assess bisulfite conversion rate after sequencing) and 5% PhiX spike-in (to improve sequencing metrics and offset the lower-complexity WGBS libraries). The WGBS libraries were sequenced on an Illumina X Ten platform with 2×150bp paired end reads.

### RNAseq data generation

Extracted RNA was subjected to QC and library construction as previously described ^17^ Briefly, libraries were constructed using the TruSeq Stranded Total RNA Library Preparation kit with Ribo-Zero Gold ribosomal RNA depletion, with ERCC Mix 1 spike-ins added to each sample. These paired-end, strand-specific libraries were sequenced on an Illumina HiSeq 3000 using 2×100bp reads.

### WGBS Data Processing

The raw WGBS data were processed using FastQC to control for quality of reads, Trim Galore to trim reads and remove adapter content ^19^, Arioc for alignment to the GRCh38.p12 genome (obtained from ftp://ftp.ncbi.nlm.nih.gov/genomes/all/GCA/000/001/405/GCA_000001405.27_GRCh38.p12/GCA_000001405.27_GRCh38.p12_assembly_structure/Primary_Assembly/assembled_chromosomes/) ^20^, duplicate alignments were removed with SAMBLASTER ^21^, and filtered with samtools ^22^ (v1.9) to exclude all but primary alignments with a MAPQ >= 5. We used the Bismark methylation extractor to extract methylation data from aligned, filtered reads ^23^. We then used the bsseq R/Bioconductor package (v1.22) to process and combine the DNA methylation proportions across the samples for all further manipulation and analysis ^24^. After initial data metrics were calculated, the methylation data for each sample was locally smoothed using BSmooth with default parameters for downstream analyses. CpG results were filtered to those not in blacklist regions (N = 27,812,354). CpHs were filtered to sites which had >3 coverage and non-zero methylation in at least half the samples. Due to an unidentifiable primary source of variance, 11 samples in the DLPFC were dropped before analysis. We also extracted DNA sequence variants from 740 common exonic/coding sites for comparisons to DNA genotyping data to confirm sample identities, as implemented in our SPEAQeasy RNA-seq software^25^.

### RNAseq Data Processing

RNA sequencing reads were processed as described in Benjamin et al using the SPEAQeasy pipeline described in Eagles et al ^25^. Briefly, reads were aligned to the human genome using HISAT2 ^26^ and genes were quantified in a strand-specific manner using featureCounts ^27^. RNA-called coding variants were used to confirm sample identities against corresponding DNA genotyping data. Exonic sequences that were susceptible to RNA degradation from an independent tissue degradation experiment were extracted from coverage-level data following our qSVA algorithm ^28^, as described in more detail in Benjamin et al.

### Differential gene expression analysis

For all differential gene expression analyses, we first filtered to samples which had exonic mapping rate > 0.37, mitochondrial mapping rate < 0.1, and RNA integrity number (RIN) > 6. We then filtered out lowly expressed genes by calculating reads per kilobase per million (RPKM) genes assigned during counting, and retaining those genes which had RPKM > 0.1. We then performed differential expression analysis across various sample subsets for the diagnosis, antipsychotic, and antipsychotic generation variables. For each variable, we performed linear modelling with limma, modelling voom-normalized feature counts (on the log2 scale) across the variable of interest, adjusting for potential confounders including age, sex, mitochondrial mapping rate, rRNA rate, exonic mapping rate, RIN, overall mapping rate, ERCC bias factor, and the top 5 quantitative ancestry factors. We further adjusted for quality surrogate variables (qSVs), which were calculated from the top k principal components (PCs) of degradation-susceptible exonic regions. We selected k = 16 using the BE algorithm with the sva Bioconductor package. While this qSVA was designed to reduce spurious differential expression signal due to RNA quality differences between groups, the latent qSVs can also capture and control for potential cell type composition variation ^10^ and thus should correct for differences between samples. To analyze differences within cases (SCZDAP vs SCZD), we applied contrasts to a model which separated samples into three groups by diagnosis and antipsychotic use. Linear modelling effects were converted to empirical Bayes-moderated T-statistics, with corresponding p-values, and Benjamini-Hochberg-adjusted (BH-adjusted) p-values using the limma topTable function.

### Differential methylation analysis

Before analysis, CpG sites were filtered to those which are outside the ENCODE blacklist ^29^, which has been shown to have poor data quality in WGBS ^9^. CpH sites were filtered to those outside the blacklist, and those which have coverage > 3 and non-zero methylation for at least half of samples. Differential methylation analyses for diagnosis and antipsychotic use were performed using linear regression modelling accounting for sex, age, estimated neuronal composition (represented by the top principal component of methylation data), and the top 3 MDS components from genotype data. The regression analyses above were formed using limma, which employed empirical Bayes and returned moderated T-statistics, which were used to calculate *P* values and estimate the false discovery rate (FDR, via Benjamini-Hochberg approach).

### Cell type enrichment of differentially expressed genes

Cell-type specific gene expression data was taken from Tran et al. ^18^ for the DLPFC and nucleus accumbens (a region which is also in the striatum with the caudate nucleus). We selected the top 2000 most cell type-specific genes for each considered cell type, which makes analyses across cell types more comparable (as many cell types had far more than 2000 DEGs that were significant). We then performed one Fisher’s exact test per cell type to assess the enrichment of subsequent cell type-specific genes against our significant antipsychotic-differentially expressed genes. This involved comparing the proportion of differentially expressed genes which were within the cell type specific gene sets to the proportion of differentially expressed genes which were not within these cell type specific gene sets.

### Comparison to mouse

To compare our results to the significantly differential genes previously reported by Kim and colleagues ^12^ in mouse, we first filtered all results to those which have mouse human orthologs (using biomaRt getLDS). We identified which genes were significant in both mouse and human analysis, and then performed enrichment analysis using Fisher’s exact test to compare how many genes were significantly differentially expressed in human and how many genes were in the significant set of mouse genes with human orthologs.

## Data Availability

All data produced in the present study will be posted online upon publication.

## Acknowledgements

The authors would like to express their gratitude to our colleagues whose tireless efforts have led to the donation of postmortem tissue to advance these studies: the Office of the Chief Medical Examiner of the District of Columbia; the Office of the Chief Medical Examiner for Northern Virginia, Fairfax Virginia; and the Office of the Chief Medical Examiner of the State of Maryland, Baltimore, Maryland. We would also like to acknowledge Llewellyn B. Bigelow, MD, for his diagnostic expertise. This project was supported by The Lieber Institute for Brain Development and by NIH grants R01MH112751 and T32GM781437. Finally, we are indebted to the generosity of the families of the decedents, who donated the brain tissue used in these studies.

